# Thrombolysis in Acute Pulmonary Embolism: are we over – doing it?

**DOI:** 10.1101/2020.09.13.20193672

**Authors:** Refai Showkathali, Radhapriya Yalamanchi, Balasubramaniam Ramakrishnan, Abraham Oomman, Aruna Sivaprakash, Pramod Kumar

## Abstract

**Aim and Method:** We aimed to study the clinical data and outcome of patients admitted in our centre with acute pulmonary embolism (PE) over a five year period from May 2013 to April 2018. The main outcome data included were- in-hospital bleeding, in hospital RV function improvement, PAH improvement, duration of hospital stay, 30 and 90 day mortality.

**Results:** A total of 114 (69 m, 55 f) patients with the mean age of 55+/− 15 yrs were included. Patients who had involvement of central pulmonary trunk- called as “Central PE” group (n = 82) and others as “Peripheral PE” group (n = 32). There were more women in the peripheral PE group (53.1% vs 34.1%, p 0.05), while RBBB (22% vs 3.1%, p 0.02) and RV dysfunction (59.8% vs 25%, p 0.002) were noted more in the central PE group. Systemic thrombolysis was done in 53 patients (49 central, 4 peripheral), of which only 3 had hypotension and 28 patients were in the Intermediate-high risk group. The overall in-hospital, 30 day and 90 day mortality were 3.6, 13.2 and 22.8% respectively. Bleeding was significantly higher in thrombolysed group compared to the non- thrombolysed group (18.9% vs 0, p 0.0003). However, improvement in pulmonary hypertension was noted more in thrombolysis group compared to non-thrombolytic group. (49% vs 21.2%, p 0.01).

**Conclusion:** This retrospective data from a tertiary centre in South India showed that short and mid-term mortality of patients with PE still remains high. The high non-guideline use of thrombolysis has reflected in the increased bleeding noted in our study.

## MANUSCRIPT

### Introduction

Acute pulmonary embolism (PE) is a common, potentially life-threatening disease and is the most serious clinical presentation of Venous thrombo-embolic disorder.[1] Mortality occurs in approximately 2 to 6% of patients in hemodynamically stable PE and in 30% or more in patients with hemodynamic instability or shock.[2–4] Of note, 25% of the patients do not survive the first year after diagnosis of PE, although the majority of deaths during this time are related to underlying conditions such as cancer or chronic heart disease rather than to PE itself.[3–4]

Over the past 25 years, thrombolytic therapy has consistently demonstrated improvement in hemodynamic parameters in patients with PE.[5] Clinically, although it results in reduced mortality in patients with massive PE, thrombolytic therapy is not beneficial in unselected patients with PE.[6–7] A review of randomized trials performed before 2004 indicated that thrombolysis may be associated with a reduction in mortality or recurrent PE in high-risk patients who present with hemodynamic instability.[7] According to the European Society of Cardiology (ESC) guidelines on the diagnosis and management of acute PE published in 2014, the only current absolute indication for thrombolysis is high risk PE (i.e PE with shock or persistent hypotension.[8] In intermediate risk patients, full dose thrombolytic therapy can prevent potenitally life threatening haemodynamic decompensation, but this benefit is counterbalanced by high risk of haemorrhagic stroke or major non-intra cranial bleeding.[9] Even in the latest ESC guidelines published few months ago, thrombolysis is indicated only in high risk PE and to consider rescue thrombolysis in intermediate-high risk PE patients.[10]

We aimed to study the clinical data including management decisions of patients presenting with acute PE in our centre over a five year period and to analyse the clinical outcome of these patients to understand the “real-world” practise of management of PE in a high volume centre in South India.

### Methods

Patients who were diagnosed to have acute PE by CT pulmonary angiogram (CTPA) over a period of 5 years (May 2013 to April 2018 inclusive) in our centre were identified by the electronic healthcare database. All our hospital case records over the last 7 years were scanned and saved electronically in a database. We retrospectively analysed the case records of these patients with their respective unique identification number. Their clinical data including baseline characteristics, imaging reports (ECHO/CTPA), clinical parameters and management strategies (including thrombolysis) were recorded. Simplified PESI score was calculated for all patients as per the guidelines.[11] The study has been approved by our Institutional Ethics committee- (Ref No- IEC-CS No- AMH-008/03-19) and the procedures followed were in accordance with the ethical standards of the responsible committee on human experimentation.

#### Definitions

Central PE- PE involving the central pulmonary trunk (main pulmonary artery, Right or Left pulmonary artery)

Peripheral PE- PE involving peripheral pulmonary artery only

Hypotension is defined as systolic BP < 90 mmHg.

Tachycardia is defined as heart rate > 100/min.

Right Ventricular (RV) dysfunction: Echocardiographic criteria of RV end-diastolic diameter of > 30 mm or hypokinesia of RV free wall noted in any view or Tricuspid annular plane systolic excursion (TAPSE < 16 mm).

Pulmonary arterial hypertension (PAH): By echocardiographic criteria of Right Ventricular Systolic Pressure (RVSP)- Normal < 40 mmHg, mild PAH-40-54 mm Hg, Moderate PAH-55-69 mmHg, Severe PAH- > 70 mmHg.

#### Outcome data

The main outcome data included were- in-hospital mortality, 30 day mortality, 90 day mortality, in-hospital bleeding, duration of hospital stay, improvement in PAH, and improvement in RV function during hospital stay. PAH improvement is defined as at-least one step improvement of PAH in the echocardiogram prior to discharge, compared to the index echocardiogram. Except for 30 day and 90 day mortaility, all other outcome data were from the index admission and was obtained from the records. For 30 day and 90 day mortality, we scanned the patients follow up visit to the hospital (to any department) with the unique ID number and considered alive if they have visited the hospital. If the details were not available, patients were contacted by phone and mail to receive further information.

#### Statistics

Continuous data are presented as mean ± standard deviation and categorical outcomes are presented as percentages. Categorical outcomes were compared by means of Fisher’s exact test and permutation unpaired t test was used to compare continuous variable between two groups. A p value of 0.05 was considered to be statistically significant.

### Results

A total of 114 (69 male, 55 female) patients with the mean age of 55+/− 15 yrs were diagnosed with acute PE by CTPA during the study period. Eighty two patients were grouped as central PE and the other 32 patients as “Peripheral PE” group. The baseline characteristics of the two groups were compared in Table 1. There were more women in the peripheral PE group (53.1% vs 34.1%, p 0.05), while RBBB (22% vs 3.1%, p 0.02) and RV dysfunction(59.8% vs 25%, p 0.002) were noted more in the central PE group.

**Table 1:** Baseline Characteristics and Management Strategy of Central vs Peripheral PE patients

|  | Total (n=114)<br>n (%) | Central PE<br>(n=82)<br>n (%) | Peripheral PE<br>(n= 32)<br>n (%) | p value |
| --- | --- | --- | --- | --- |
| Age in yrs | 55+/-15 | 56 +/- 15 | 52 +/- 16 | 0.15 |
| Female | 45 (39.5) | 28 (34.1) | 17 (53.1) | 0.05 |
| DVT | 67 (58.8) | 47 (57.3) | 20 (62.5) | 0.52 |
| Hypotension | 5 (4.4) | 5 (6.1) | 0 | 0.32 |
| Tachycardia | 64 (56.1) | 48 (58.5) | 16 (50) | 0.53 |
| RBBB | 19 (16.7) | 18 (22) | 1 (3.1) | 0.02 |
| RV dysfunction | 57 (50%) | 49 (59.8) | 8 (25) | 0.002 |
| Any PAH | 72 (63.2) | 55 (67.1) | 17 (53.1) | 0.19 |
| Mild PAH | 36 (31.6) | 29 (35.4) | 7 (21.9) | 0.26 |
| Moderate PAH | 21 (18.4) | 15 (18.3) | 6 (18.8) | 1.0 |
| Severe PAH | 15 (13.2) | 11 (13.4) | 4 (12.5) | 1.0 |
| No PAH | 42 (36.8) | 27 (32.9) | 15 (46.9) | 0.19 |

A total of 53 patients were thrombolysed for PE (49 in central and 4 in peripheral PE group), of which alteplase is the most commonly used agent. (Table 2) Vitamin K antagonists was used in 81 patients and NOAC in 30 patients. Three patients died before starting any oral anticoagulants. Apixaban is the most commonly used NAOC (14.9%) compared to Dabigatran (3.5%) and Rivoroxaban (7.9%). There were no difference in outcome between the central and peripheral PE group. (Table 3)

**Table 2:** Management of patients with central and Peripheral PE.

|  | Total (n=114)<br>n (%) | Central PE<br>(n=82)<br>n (%) | Peripheral PE<br>(n= 32)<br>n (%) | p value |
| --- | --- | --- | --- | --- |
| Thrombolysed | 53 (46.5) | 49 (59.8) | 4 (12.5) | 0.0001 |
| Alteplase | 42 (36.8) | 40 (48.8) | 2 (6.3) | 0.0001 |
| Tenecteplase | 9 (7.9) | 7 (8.5) | 2 (6.3) | 1.0 |
| Streptokinase | 2 (1.8) | 2 (2.4) | 0 | 1.0 |
| IV Heparin | 38 (33.3) | 27 (32.9) | 11 (34.4) | 0.83 |
| LMWH | 105 (92.1) | 76 (92.7) | 29 (90.6) | 1.0 |
| VKA | 81 (71.1) | 57 (69.5) | 24 (75) | 0.49 |
| NOAC | 30 (26.3) | 23 (28) | 7 (21.9) | 0.64 |
| Apixaban | 17 (14.9) | 13 (15.9) | 4 (12.5) | 0.78 |
| Dabigatran | 4 (3.5) | 4 (4.9) | 0 | 0.57 |
| Rivoroxaban | 9 (7.9) | 6 (7.3) | 3 (9.4) | 0.70 |

**Table 3:** Clinical Outcome of central vs peripheral PE patients.

|  | Total<br>(n=114)<br>n (%) | Central<br>(n=82)<br>n (%) | Peripheral<br>(n= 32)<br>n (%) | p value |
| --- | --- | --- | --- | --- |
| In-hospital bleeding | 10 (8.8) | 10 (12.2) | 0 | 0.06 |
| Duration of hospital stay (days) | 7.5 +/- 3.9 | 7.3+/- 3.7 | 8.1+/-4.3 | 0.6 |
| PAH improvement | 41 (36) | 31 (37.8) | 10 (32.3) | 0.66 |
| RV dysfunction improvement | 4 (3.5) | 2 (2.4) | 2 (6.5) | 0.30 |
| In-hospital Mortality | 4 (3.6) | 4 (4.9) | 0 | 0.58 |
| 30-day mortality | 15 (13.2) | 10 (12.2) | 5 (15.6) | 0.76 |
| 90-day mortality | 26 (22.8) | 16 (19.5) | 10 (31.3) | 0.22 |

There was no significant difference in mortality between thrombolysis and non thrombolysed group (Fig 1). Bleeding was significantly higher in thrombolysed group compared to the non- thrombolysed group (18.9% vs 0, p 0.0003). There was one step improvement in PAH in the thrombolysed group (50.9% vs 23%, p 0.003).

**Fig 1:**
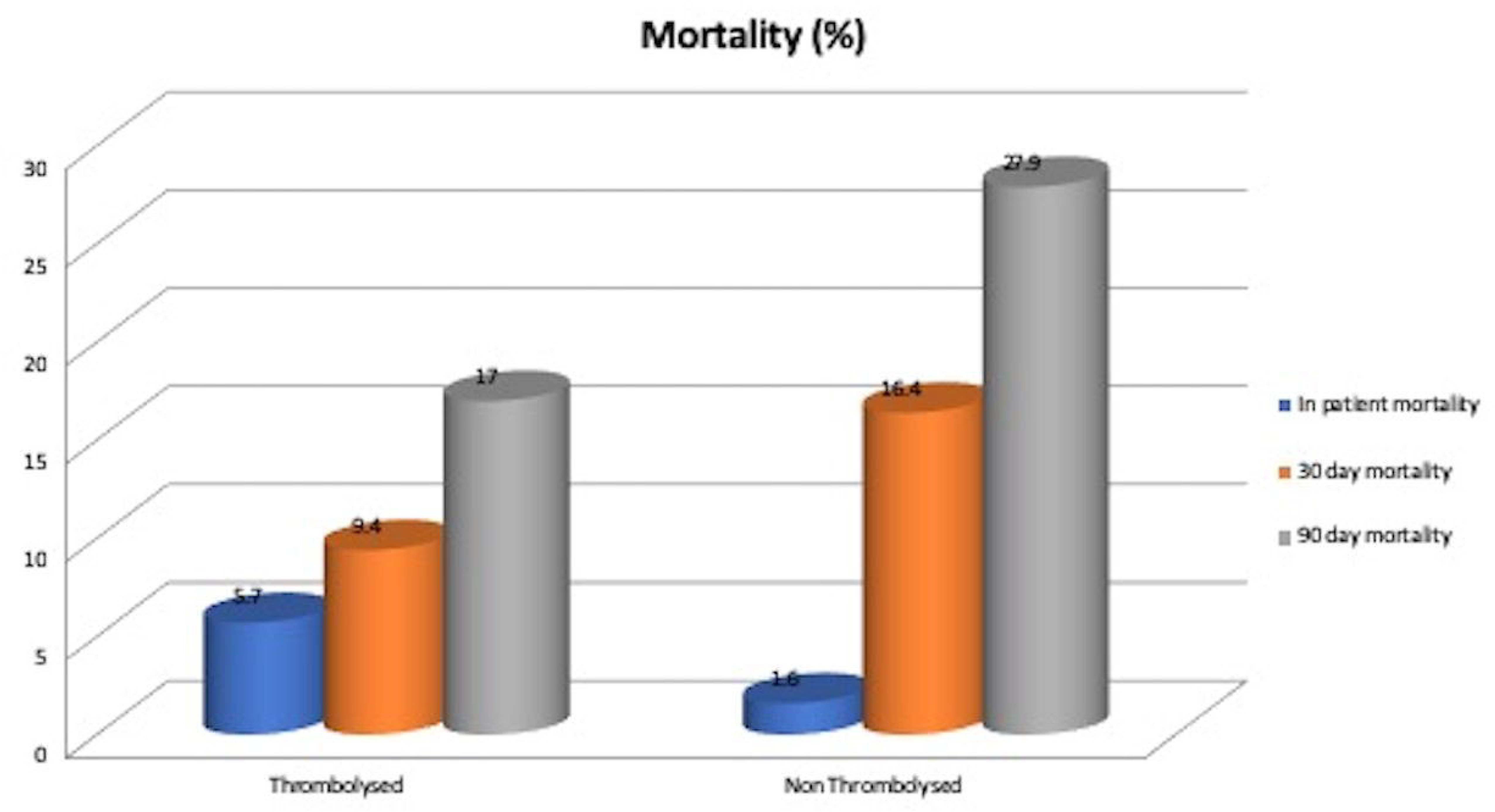
Mortality outcome of Thrombolysed vs Non-Thrombolysed patients.

Out of those patients with central PE (n = 82), 49 were thrombolysed- the indication was high risk in 3 (6.1%), Intermediate high in 28 (57.1%), but no clear indication in 18

(36.8%) patients. On comparing with central PE patients who were not thrombolysed (n = 33), thrombolysed group had more patients with any form of PAH (77.6% vs 51.5%, p 0.02) or RV dysfunction (73.5% vs 39.4%, p 0.002) (Table 4). Comparison of outcome of thrombolysis vs non-thrombolysis groups showed bleeding occurred more commonly in thrombolysed patients (20.4% vs 0, p 0.004) with no difference in mortality or duration of hospital stay (Table 5). However, PAH improvement was noted more in thrombolysis group compared to non thrombolytic group. (49% vs 21.2, p 0.01). The bleeding rate was much higher in patients who had streptokinase (50%), while patients who had alteplase and tenecteplase had 16.7% and 22.2% bleeding respectively.

**Table 4:** Clinical characteristics of Central PE patients who were Thrombolysed vs Non-thrombolysed

|  | TOTAL<br>(n= 82)<br>n (%) | Thrombolysed<br>(n= 49)<br>n (%) | Non Thrombolysed<br>(n= 33)<br>n (%) | P value |
| --- | --- | --- | --- | --- |
| Age | 55.9+/-15.1 | 55.8+/-13.5 | 55.9 +/-17.5 | 0.91 |
| Female | 28 (34.1) | 17 (34.7) | 11 (33.3) | 1.0 |
| RBBB | 18 (22) | 14 (28.6) | 4 (12.1) | 0.1 |
| Any PAH | 55 (67.1) | 38 (77.6) | 17 (51.5) | 0.02 |
| RV dysfunction | 49 (60) | 36 (73.5) | 13 (39.4) | 0.002 |
| Hypotension | 5 (6.1) | 3 (6.1) | 2 (6.1) | 1.0 |
| sPESI $\geq 1$ | 56 (68.3) | 44 (89.8) | 12 (36.3) | 0.0001 |
| Positive Troponin | 42 (51.2) | 30 (61.2) | 12 (36.3) | 0.04 |
| Intermediate high risk | 34 (41.4) | 28 (57.1) | 6 (18.2) | 0.0006 |

**Table 5:** Outcome differences of Central PE patients who were thrombolysed vs not thrombolysed

|  | TOTAL<br>(n= 82)<br>n (%) | Thrombolysed<br>(n= 49)<br>n (%) | Non<br>Thrombolysed<br>(n= 33)<br>n (%) | P value |
| --- | --- | --- | --- | --- |
| In-hospital<br>Bleeding | 10 (12.2) | 10 (20.4) | 0 | 0.004 |
| RV dysfunction<br>improvement | 2 (2.4) | 1 (2.0) | 1 (3.03) | 1.0 |
| PAH<br>improvement | 31 (37.8) | 24 (49.0) | 7 (21.2) | 0.01 |
| Duration of<br>hospital stay in<br>days (mean±SD) | 7.3+/- 3.7 | 7+/- 3 | 7.9+/-4.6 | 0.46 |
| In-hospital<br>Mortality | 4 (4.9) | 3 (6.1) | 1 (3.0) | 0.64 |
| 30 day mortality | 10 (12.2) | 5 (10.2) | 5 (15.2) | 0.51 |
| 90- day mortality | 16 (19.5) | 8 (16.3) | 8 (24.2) | 0.41 |

Two patients who had central PE and hypotension were not thrombolysed. (one due to previous Intra cranial haemorrhage (ICH) and other due to unknown reason). Four patients in the peripheral PE group who got thrombolysed were done in the first year of the study (2013-2014), before proper guidelines were released.

Ten patients who were thrombolysed had some form of bleeding- 4 had gastric, 2 had rectal, one had ICH, one had haemoptysis, one had gum and another one had conjunctival bleeding. Three patients who had gastric bleed and one who had rectal bleed needed RBC transfusion. One patient who had gastric bleed died while in-hospital and another patient who had gastric bleed died within 90 days. The other 8 patients were alive until 90 days of follow up.

### Discussion

This “real-world” study of patients with PE in a high volume centre, suggest that thrombolysis were more commonly used than guideline advised indications. In 18 patients in the central PE group who had thrombolysis (5 in the year 2013, 5 in 2014, 4 in 2015, 3 in 2016 and 1 in 2017), there was no clear indication identified from the medical notes of the patient for initiating thrombolysis. In general there was a low threshold for giving thrombolytic treatment, particularly for patients with central PE, adapted by most clinicians and hospitals until recent years. Even though there was no significant mortality noted in this retrospective data from a high volume centre in patients who were thrombolysed, there was an increased risk of bleeding with thrombolytic therapy.

The latest ESC and the American College of Chest Physicians guidelines recommend thrombolysis only for those patients with clinical signs of haemodynamic decompensation.[8,10,12–13] The ESC, for example, classifies thrombolytic administration in patients with acute high-risk PE as a 1B recommendation, and the 2016 updated CHEST guidelines list it as a grade 2B recommendation.[8,12] The guidelines for thrombolysis in high risk PE patient comes from randomised trials. A large meta analysis done in 2004 showed there were benefits in thrombolysing high risk PE patients.[7]

There has been always a controversy about the use of thrombolytic therapy in intermediate risk patients until PEITHO trial was published.[14] PEITHO trial is a large randomised study which compared the outcome of intermediate risk PE patients with or without thrombolysis. In this study, thrombolysis with tenecteplase showed significant reduction in the risk of haemodynamic decompensation within 7 days. However, thrombolysis was also associated with a 10- fold increase in intracranial haemorrhage (2% vs 0.2%) and a five-fold increase in major haemorrhage (6.3% vs 1.2%).[9] The follow-up results of the same study showed that thrombolysis with tenecteplase in intermediate risk PE patients did not affect the long term survival.[14] Despite this study publication in 2017, Tenecteplase is not approved by FDA and ESC for usage in PE.

The ESC 2014 and 2019 guidelines recommends clinical risk assessment of those PE patients without hypotension by using PESI score, to further stratify the management strategy. Patients who have a sPESI ≥ 1 are considered intermediate risk and further divided into Intermediate-high risk and Intermediate-low risk depending on RV function and labarotary tests like natriuretic peptides and Troponin. Those Intermediate-high risk patients can also be considered for rescue reperfusion therapy with thrombolytic agents. There are no other indication for thrombolysis in PE according to these guidelines. Even in the latest published retrospective study from a single centre in US, only 15 out of 196 (7.6%) patients had thrombolytic therapy.[15] Out of the 15 patients, 4 are considered high risk and the other 11 were considered to be intermediate risk according to PESI score. Alteplase is the only agent used in their study, as that is the only FDA approved thrombolytic therapy for PE in US. Major extracranial bleeding occurred in 12 patients (6.1%) of the whole cohort in their study, but interestingly only 2 out of 15 patients (13.3%) who were thrombolysed had bleeding. The other 10 patients who had major extracranial bleed did not underwent thrombolysis.[15] In our study, thrombolysis rates were much higher at 46.5%. Out of the 53 patients who were thrombolysed, alteplase was used in 42 (79.2%), tenecteplase in 9 (16.9%) and streptokinase in 2 (3.7%) patients. Some form of bleeding occurred in 10 patients in our study, but all these 10 patients had thrombolytic therapy (18.9%), with zero bleeding in the non thrombolytic group.

### Limitations

We included only patients who had confirmed PE on CTPA. Patients with massive PE sometimes can present with sudden cardiac arrest with no time to undergo CTPA (either died or had thrombolysis with echocardiographic findings). This particular group of patients were not included in our study. As this was a retrospective study, we did not have the accurate data about these patients and therefore not included.

### Conclusion

This retrospective data from a tertiary centre in South India showed that short and mid term mortality of patients with PE remains high despite early diagnosis and management. This study has shown that there was increased usage of thrombolytic therapy, even in those patients who did not fulfil the criteria for thrombolysis. This has led to the higher incidence of bleeding, even though some of them are non-life threatening bleeds. Clinicians should be aware of the indications for thrombolysis in PE and to risk stratify them accordingly in their day to day clinical practise.

## Data Availability

Yes. Available in our database

